# Interaction between placental efflux transporters and use of antiseizure or antidepressant drugs during pregnancy on birth weight in the Norwegian Mother, Father and Child Cohort Study

**DOI:** 10.1101/2023.09.13.23295417

**Authors:** Marta H. Hernandez, Jacqueline M. Cohen, Karoline H. Skåra, Thea K. Grindstad, Yunsung Lee, Per Magnus, Pål R. Njølstad, Ole A. Andreassen, Elizabeth C. Corfield, Alexandra Havdahl, Espen Molden, Kari Furu, Maria C. Magnus, Alvaro Hernaez

## Abstract

**Background:** Intrauterine exposure to antiseizure and antidepressant drugs is associated with adverse pregnancy outcomes, including low birth weight. Gene variants in placental efflux transporter genes may alter foetal exposure to these drugs.

**Methods:** We investigated whether genetic variants in placental efflux transporters modified the impact of maternal antiseizure and antidepressant drug use on offspring birth weight, using data from the Norwegian Mother, Father and Child Cohort Study and the Medical Birth Registry of Norway (69,828 offspring with their genotype data, 81,189 with maternal genotype data). We systematically searched for gene variants in placental efflux transporters influencing drug exposure [MDR1-*ABCB1* (7 alleles), MRP1-*ABCC1* (2), MRP2-ABCC2 (3) and, BCRP-*ABCG2* (2)] and calculated genetic scores (sum of alleles potentially related to low transporter activity). We assessed the interaction between prenatal drug use and genetic scores on birth weight.

**Findings:** Prenatal antiseizure medication exposure was associated with lower birth weight (-95.5 grams, 95% CI -190 to -0.78). This relationship depended on the offspring’s MRP2-*ABCC2* genetic score. Birth weight differences between exposed vs. unexposed were 70.3 g (95% CI -494 to 634) in the lowest genetic score category and -306 g (95% CI -361 to -31.8) in the highest (interaction *p*-value = 0.019; main variant: rs3740066). The antiseizure medication-lower birth weight association also depended on the maternal MDR1-*ABCB1* genetic score. Birth weight differences between exposed vs. unexposed were -66.8 g (95% CI -225 to 91.2) in the lowest genetic score category and -317 g (95% CI -517 to -117) in the highest (interaction *p*-value = 0.037; main variants: rs10248420 and rs2235015). Prenatal antidepressant exposure was associated with low birth weight, but no gene-drug interactions were observed.

**Conclusions:** MRP2-*ABCC2* and MDR1-*ABCB1* variants may influence prenatal antiseizure medication’s impact on neonatal birth weight.

**SUMMARY:** *What is already known on this topic:*

- Low birth weight increases the risk of neonatal death, morbidity, and lifelong susceptibility to various health conditions, including neurodevelopmental, cardiometabolic, respiratory, and psychiatric disorders.
- Previous studies have reported an association between maternal use of centrally acting medication during pregnancy and low birth weight in the offspring, but the underlying causes remain poorly understood.
- It is of great concern to identify strategies that can modulate foetal exposure to these medications and enable individualised, safe use since discontinuation may also increase the risk of harm to the mother.

*What this study adds:*

- Our study uncovered genetic variants in both offspring (rs3740066) and mother (rs10248420 and rs2235015) linked to placental efflux transporters that modulated the association between maternal antiseizure drug use during pregnancy and the risk of low birth weight in the offspring, marking the first exploration of such interactions.
- If these findings are confirmed and information about specific drugs is incorporated, this could potentially be used in the development of personalized recommendations for pregnant women undergoing these treatments.

## INTRODUCTION

The use of medication during pregnancy must carefully balance benefits to the mother and potential harm to the offspring.^1^ Centrally acting drugs (such as antiseizure drugs [mainly lamotrigine, 0.3% of pregnancies] and antidepressants [1.5% of pregnancies]) have been increasingly used during pregnancy.^2,3^ However, intrauterine exposure to these medications may be associated with adverse pregnancy outcomes such as impaired foetal growth and reduced birth weight in offspring.^4–6^ Furthermore, these conditions are linked to a multitude of morbidities, resulting in both immediate and long-term adverse consequences for newborns, their families, and society.^7^ The placenta plays an essential role in substance exchange between mother and foetus (e.g., nutrients, metabolic by-products). Small, non-polar, lipophilic xenobiotics such as antiseizure drugs and antidepressants can pass through the placenta via passive diffusion.^8^ However, several active proteins can efflux these compounds from the foetus back to the mother.^9^ Active efflux proteins from the adenosine triphosphate-binding cassette superfamily (such as P-glycoprotein, breast cancer resistance protein, and others multidrug resistance proteins) are involved in the transplacental passage of drugs particularly affecting the passage of centrally-acting drugs and their metabolites.^10,11^ Several antiseizure drugs and antidepressants are substrates for these transporters.^12,13^ Efflux transporters may therefore have a foetoprotective effect by lowering the concentrations of potentially toxic drugs on the foetal side. Genetic variants in the transporters may modify their activity and could be located in the offspring or maternal genotype. ^14,15^ Little is known about the role of genetic variants in placental efflux transporters on the modulation of the association of antiseizure drugs and antidepressants during pregnancy with offspring outcomes such as low birth weight.

Our aim was to explore whether there is an interaction between the use of antiseizure drugs or antidepressants during pregnancy and genetic variants related to placental efflux transporters (in P-glycoprotein, breast cancer resistance protein, and others multidrug resistance proteins) on offspring birth weight.

## MATERIALS AND METHODS

### Study participants

We included participants in the Norwegian Mother, Father, and Child Cohort Study (MoBa). MoBa is a prospective, population-based pregnancy cohort conducted by the Norwegian Institute of Public Health. Pregnant women and their partners were recruited across Norway between 1999-2008 at the time of routine ultrasound screening (∼17^th^ gestational week). The cohort includes approximately 114,000 children, 95,000 mothers, and 75,000 fathers.

This work used a subsample of offspring from singleton pregnancies with available information on genotype and birth weight (from version #12 of the quality-assured data files released on May 11, 2022). Genotype data was obtained from blood samples provided during pregnancy and at birth (mothers) and umbilical cord blood at birth (offspring).^16^ A total of 238,001 samples have been genotyped in 24 genotyping batches with varying selection criteria, genotyping platforms, and genotyping centres.^17^ Quality control, phasing, and imputation were performed using the MoBaPsychGen pipeline as previously described.^17^

Our work is described according to the Strengthening the Reporting of Observational studies in Epidemiology guidelines (Supplemental Table 3).^18^

### Use of centrally acting drugs

We used self-reported information on the use of centrally acting drugs from MoBa questionnaires in the 18^th^ and 30^th^ gestational weeks.^19^ Specifically, women were asked whether they had epilepsy or depression, and if they answered yes, whether any medications were used to treat the condition(s). This project has no information on the specific drug(s) the woman used. Any reported use of medication(s) for epilepsy (yes/no) or depression (yes/no) were categorized and evaluated separately.

### Birth weight

Data on offspring birth weight in grams was obtained from the Medical Birth Registry of Norway, a national health registry which contains information about all births in Norway since 1967.^20^

### Covariates

Information on maternal age at delivery (continuous), parity (1, 2, 3, ≥ 4) and sex of the offspring (female/male) was obtained from the birth registry. Furthermore, information on maternal years of education (continuous), pre-pregnancy body mass index (continuous), having ever smoked (yes/no), and self-reported depression or epilepsy was gathered in the questionnaire administered at gestational week 18.

### Genetic variants related to placental transporters and genetic scores

We performed a systematic review in PubMed, Web of Science, pharmacogenetic websites (https://www.pharmgkb.org) and GWAS catalog (https://www.ebi.ac.uk/gwas/) to identify gene variants on the adenosine triphosphate-binding cassette superfamily of placental efflux transporters (MDR1-*ABCB1*, MRP1-*ABCC1,* MRP2-*ABCC2*, and BCRP-*ABCG2*). We followed the recommendations of the Preferred Reporting Items for Systematic reviews and Meta-Analyses statement.^21^ A detailed description of the search is available in Supplemental Materials.

After the systematic review, we only conserved genetic variants that were not in linkage disequilibrium according to linkage disequilibrium block analysis in the 1000 Genomes CEU and GBR populations (given the MoBa participants passing the post-imputation quality control cluster with these populations; *R*^2^ < 0.8) and with a minor allele frequency > 1%.^22^ Genetic scores were calculated as the sum of the number of risk alleles for all genetic variants related to each transporter in a participant. The genetic score was represented in the results as a genetic score if the number of score values were ≤5. Otherwise, we represented them in quartiles. We considered a risk allele to be the allele associated with any of these five theoretical situations potentially related to an increased exposure of the drug to the foetus: decreased function of the transporter, declined expression of the transporter, non-resistance to the treatment, higher concentrations of the drug, or more adverse effects. Information related to the studies is available in Supplemental Table 2.

### Ethical approval

The MoBa study is conducted according to the Declaration of Helsinki for medical research involving human subjects. The establishment of MoBa and initial data collection was based on a license from the Norwegian Data Protection Agency. It is now based on regulations related to the Norwegian Health Registry Act. Participants provided written informed consent before joining the cohort. This project was approved by the Regional Committee for Medical and Health Research Ethics of South/East Norway (reference: 2017/1362).

### Statistical analyses

Normally distributed continuous variables were described by means and standard deviations, nonnormally distributed continuous variables by medians and 25^th^-75^th^ percentiles, and categorical variables by proportions.

We first assessed the relationship between maternal use of centrally acting drugs during pregnancy and birth weight using multivariable linear regressions adjusted for offspring sex and maternal factors (age at delivery, years of education, pre-pregnancy body mass index, parity and having ever smoked). Clustered standard errors were computed to account for dependence among births to mothers contributing with more than one pregnancy in MoBa. To determine whether there were significant interactions between maternal prenatal use of centrally acting drugs and the genetic scores on the offspring birth weight, we applied likelihood ratio tests between nested linear regression models with and without an interaction product-term of “exposure group × genetic score”. The nested models were further adjusted for the first twenty ancestry-informative genetic principal components and genotyping batch. We considered any interaction with a *p*-value for the overall interaction test < 0.05 and a differential association between medication use and offspring birth weight in the two extreme categories according to the genetic score. Whether an interaction was found, we further explored interactions with the individual variants in the genetic score as sensitivity analyses using the same strategy (as these were exploratory sensitivity analyses, we here considered any interaction with a *p*-value < 0.1).

Statistical analyses were performed in R Software, version 4.1.0. Our analysis code is available in GitHub: https://github.com/alvarohernaez/Gene_psychoactivedrug_BW_MoBa/.

## RESULTS

### Study population

In our study, we included 69,828 singleton pregnancies with offspring genotype information and offspring birth weight, and 81,189 singleton pregnancies with maternal genotype data and offspring birth weight (Figure 1). Mean birth weight was 3,639 grams (SD 522). A total of 174 children (0.25%) were exposed to maternal use of antiseizure drugs during pregnancy and 766 (1.10%) to antidepressants. Women using centrally acting drugs were younger (only for antiseizure drugs), had lower educational attainment, and were more likely to have ever smoked (Table 1).

**Figure 1.**
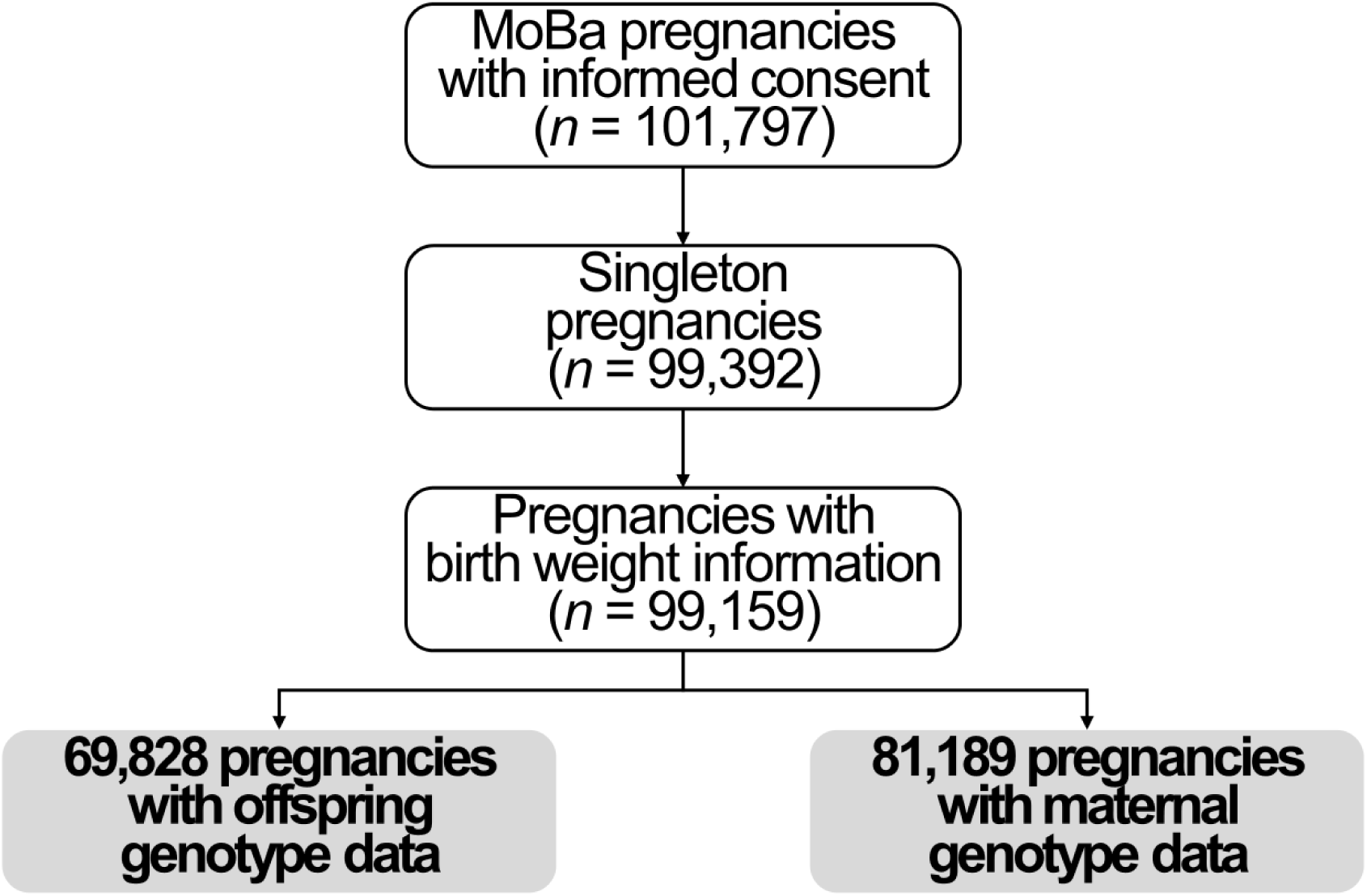
Flow chart of study participants.

**Table 1.** Baseline characteristics of mothers with offspring genotype information.

|  | **Antiseizure drugs** |  | **Antidepressants** |  |
| --- | --- | --- | --- | --- |
|  | **Exposed**<br>**(*n* = 174)** | **Unexposed**<br>**(*n* = 69,654)** | **Exposed**<br>**(*n* = 766)** | **Unexposed**<br>**(*n* = 69,062)** |
| Age at delivery (years), mean ± SD | 29.1 ± 4.83 | 30.2 ± 4.52 | 30.2 ± 5.07 | 30.2 ± 4.51 |
| Education years, mean ± SD | 15.9 ± 3.64 | 17.1 ± 3.30 | 16.2 ± 3.63 | 17.2 ± 3.30 |
| Pre-pregnancy body mass index (kg/m<sup>2</sup>), median (25<sup>th</sup>-75<sup>th</sup> percentiles) | 23.3 (21.6-26.4) | 23.3 (21.1-25.9) | 23.2 (21.1-26.6) | 23.1 (21.1-25.9) |
| Having ever smoked, *n* (%) | 107 (61.8%) | 34,310 (50.0%) | 510 (67.4%) | 33,907 (49.9%) |
| Previous number of deliveries: |  |  |  |  |
| 0, *n* (%) | 64 (36.8%) | 23,589 (33.9%) | 261 (34.1%) | 23,343 (33.9%) |
| 1, *n* (%) | 42 (24.1%) | 22,342 (32.1%) | 223 (29.1%) | 22,113 (32.1%) |
| 2, *n* (%) | 44 (25.3%) | 13,697 (19.7%) | 156 (20.4%) | 13,547 (19.7%) |
| 3, *n* (%) | 15 (8.62%) | 6,118 (8.78%) | 72 (9.40%) | 6,045 (8.77%) |
| ≥4, *n* (%) | 9 (5.17%) | 3,908 (5.61%) | 54 (7.05%) | 3,851 (5.59%) |
| Offspring birth weight, mean ± SD | 3,516 ± 603 | 3,639 ± 522 | 3,574 ± 512 | 3,640 ± 522 |
| Biological sex of the offspring: |  |  |  |  |
| Female, *n* (%) | 84 (48.3%) | 35,549 (51.0%) | 366 (47.8%) | 35,267 (51.1%) |
| Male, *n* (%) | 90 (51.7%) | 34,105 (49.0%) | 400 (52.2%) | 33,795 (48.9%) |

### Search of genetic variants on placental transporters

The systematic search identified 26 genetic variants in the placental transporter genes associated with differences in antiseizure drugs and/or antidepressant outcomes (Supplemental Figure 1), of which 14 were independent and available in the MoBa genotype database (Supplemental Table 1). Seven variants were found for MDR1-*ABCB1*, two for MRP1-*ABCC1,* three for MRP2-*ABCC2*, and two for BCRP-*ABCG2*. We used these genetic variants to calculate risk scores. The ranges of values for the four genetic scores calculated were: 0 to 11 (MDR1-*ABCB1*; we presented the stratified associations according to quartiles), 0 to 4 (MRP1-*ABCC1*), 2 to 6 (MRP2-*ABCC2*), and 0 to 4 (BCRP-*ABCG2*).

### Interaction between genetic variants of placental transporters and exposure to antiseizure drugs during pregnancy on birth weight

Antiseizure medication use during pregnancy was associated with lower birth weight (-95.5 g, 95% confidence interval [CI] -190 to -0.78). Greater values of the MRP2-*ABCC2* genetic score in the offspring were associated with lower birth weight in offspring of mothers who used antiseizure drugs during pregnancy. The difference in birth weight between exposed vs. unexposed was 70.3 g (95% CI -494 to 634) in the lowest genetic score category (2 risk alleles) and -306 g (95% CI -361 to -31.8) in the highest (6 risk alleles) (*p*-value for interaction = 0.019; Figure 2). The variant with the strongest evidence of an interaction in the sensitivity analyses of the MRP2-*ABCC2* genetic score was rs3740066 (*p*-value for interaction = 0.023; Supplemental Figure 2).

**Figure 2.**
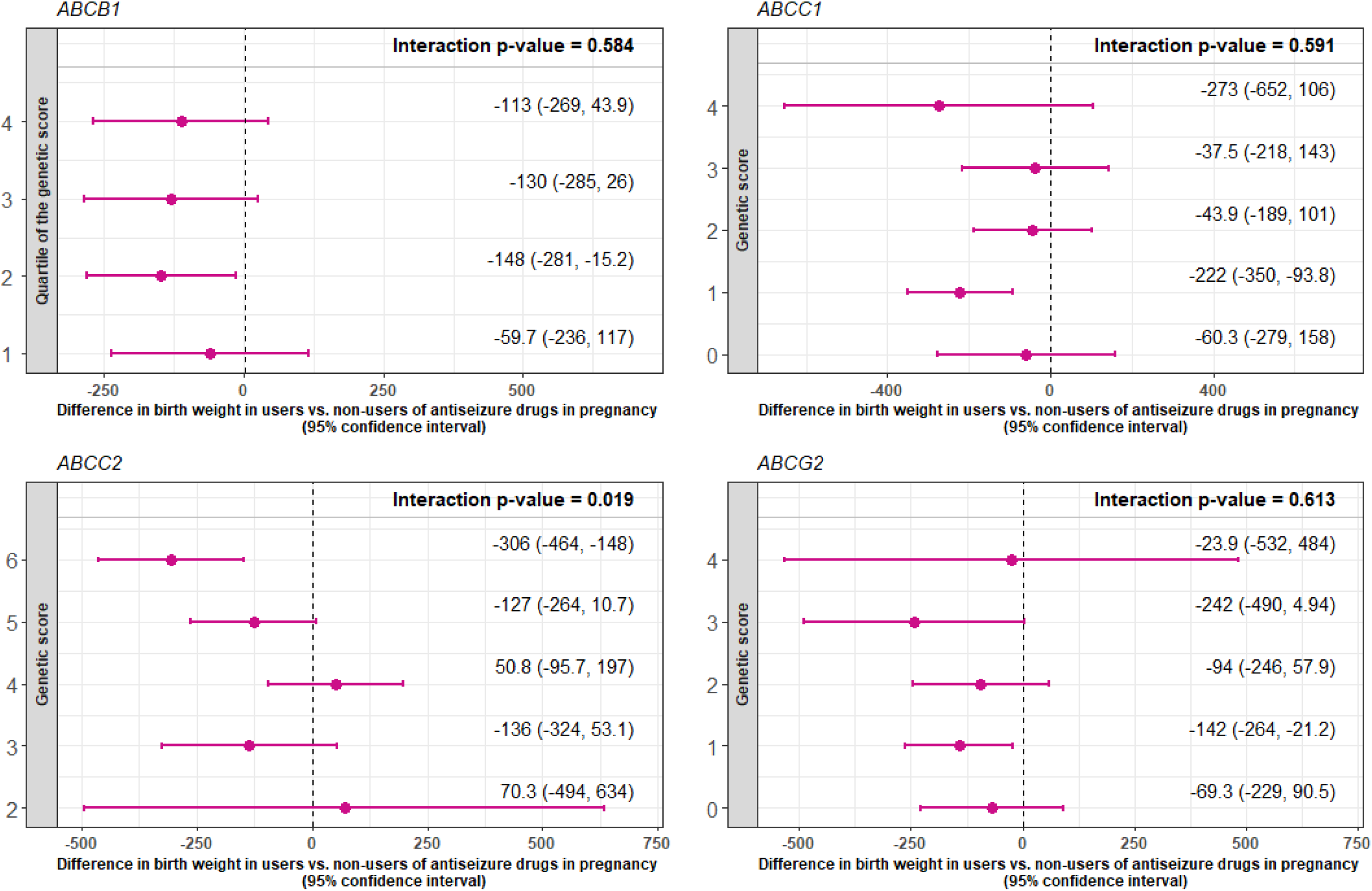
Differences in birth weight associated with pregnancy exposure to antiseizure medication due to the genetic scores for placental transporters in the offspring.

Greater values of the MDRP1-*ABCB1* genetic score in the mother were also linked to lower birth weight among offspring of mothers who used antiseizure drugs during pregnancy. The difference in birth weight between exposed vs. unexposed were -66.8 g (95% CI -225 to 91.2) in the lowest genetic score category (first quartile) and - 317 g (95% CI -517 to -117) in the highest (fourth quartile) (*p*-value for interaction = 0.037; Figure 3). Variants rs10248420 and rs2235015 included in the MDRP1-*ABCB1* genetic score had some evidence of an interaction in the sensitivity analyses (*p*-values for interactions = 0.042 and 0.058, respectively; Supplemental Figure 3).

**Figure 3.**
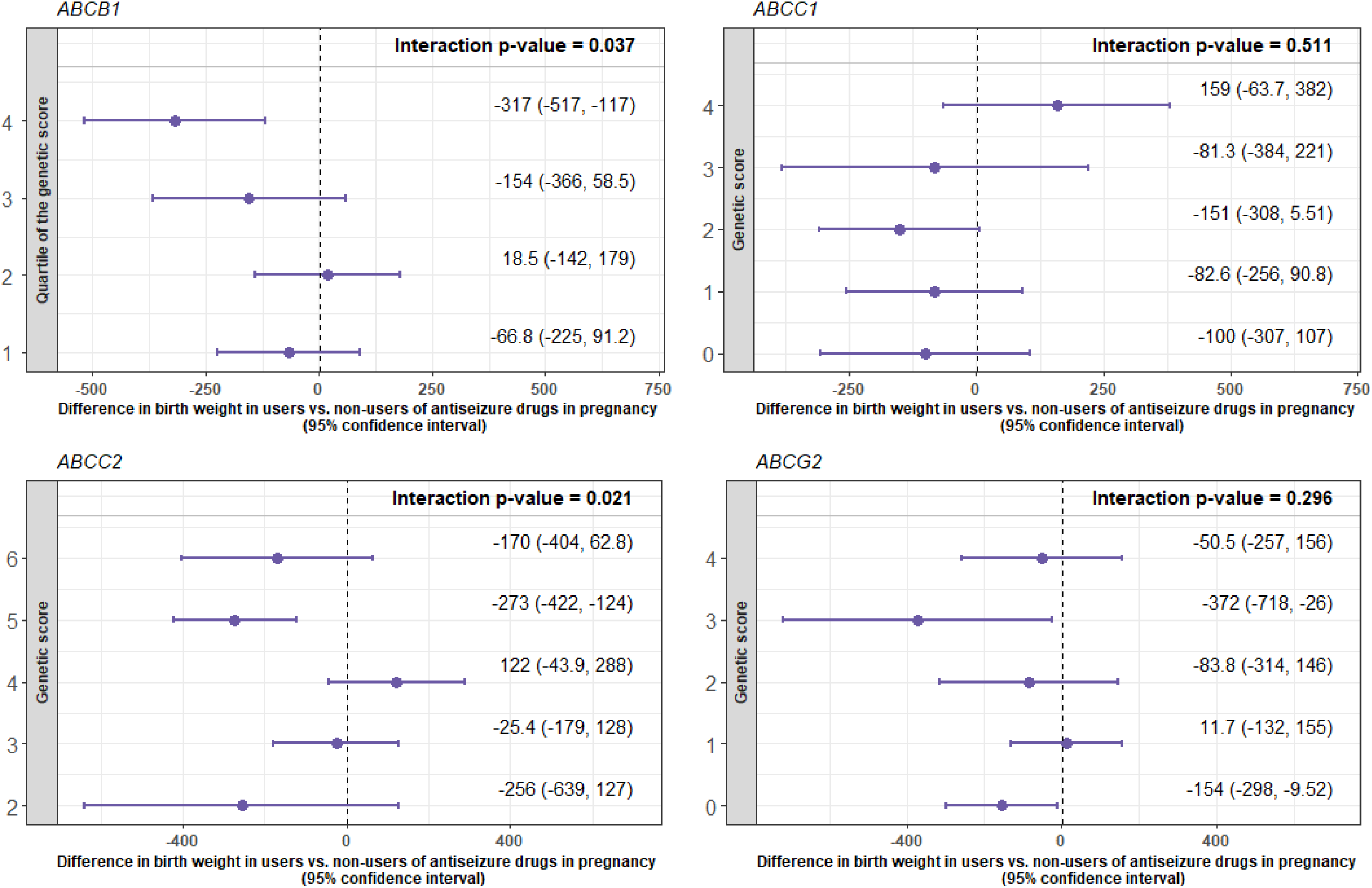
Differences in birth weight associated with pregnancy exposure of antiseizure medication due to the genetic scores for placental transporters in the mother.

### Interaction between genetic variants of placental transporters and exposure to antidepressants during pregnancy on birth weight

Use of antidepressant drugs during pregnancy was associated with lower birth weight (-60.5 g, 95% CI -97.6 to -23.3). However, we observed no interactions between prenatal use of antidepressants and any genetic score of placental efflux transporters (in offspring or mothers) on birth weight in the offspring (Supplemental Figures 4 and 5).

## DISCUSSION

In this large population-based study, involving offspring of mothers who used antiseizure medication during pregnancy, a high genetic score for MRP2-*ABCC2* in the offspring and MDR1-*ABCB1* in the mother, suggesting a greater exposure to these drugs, was associated with lower birth weight in the offspring.

MDR1-*ABCB1* and MRP2-*ABCC2* are the only transporters located in the apical membrane of the trophoblast (maternal side) and their function is to return xenobiotics to maternal circulation.^23^ Thus, it seems plausible that genetic variants linked to lower transporter activity are associated with higher foetal exposure and greater toxicity of antiseizure drugs. In addition, the main variant in the interaction between the MRP2-*ABCC2* genetic score and prenatal antiseizure drug use (rs3740066) has been previously linked to a poorer response to antiseizure medication^24^ and a higher risk of other adverse events during pregnancy (intrahepatic cholestasis of pregnancy).^25^ The differences in the interactions found within genetic variants in the offspring (MRP2-*ABCC2*) and maternal genome (MDR1-*ABCB1*) can be explained by physiological changes in the placenta structure. Early in pregnancy, the separation between the maternal circulation and the foetal circulation by the trophoblasts villous barrier is 50-100 μm (second month). It becomes progressively thinner as the pregnancy proceeds (only 4-5 μm at term).^26^ In addition, the total placenta surface area increases from about 5 m^2^ at 28 weeks of gestation to 12 m^2^ at term. Thus, passive diffusion of medications increases with gestational age, which means that medication can reach the foetal circulation more easily later in pregnancy.^26^ At the same time, the expression of MRP2-*ABCC2* increases with advancing gestational age, whereas the expression of MDR1-*ABCB1* declines.^27,28^ As a consequence of all these phenomena, the presence of risk alleles for MRP2-*ABCC2* may play a more significant role in determining the offspring’s exposure. However, an alternative hypothesis could also explain our findings. A greater foetal exposure to medication due to lower activity of the efflux transporters may also be linked to lower medication levels in the mother and a poorer control of the disease. Greater disease effects in mothers may be associated with alternative mechanisms that may also explain an increased risk of low birth weight in the offspring.

There could be several reasons for the lack of gene-drug interactions with antidepressants. First, the association between exposure to antidepressants during pregnancy and birth weight is of a smaller magnitude in our data, which can make the search of gene-drug interactions more challenging. Second, the accuracy of our data regarding the usage of antidepressants may be less precise, as women may have varying criteria for identifying such treatments. This discrepancy may not be present regarding antiseizure drugs. Finally, a larger proportion of women may discontinue the use of antidepressants during pregnancy compared to antiseizure medications.^29^

Our study has some limitations. First, due to the unavailability of information on the specific drug utilized, we conducted our analysis based on aggregated drug groups. Although the main medications in each group show similar pharmacodynamic behaviour and their usage during pregnancy reduces the number of medications under consideration,^3^ this may interfere with our findings because there may be variations in the medication groups’ interactions due to differences in transporter affinity or dose-dependent effects.^26,30^ Second, due to the lack of a genome-wide association study showing which genetic variants are associated with high or low activity of efflux transporters, we calculated risk scores using studies on genetic variants indirectly related to high or low activity (e.g., treatment resistance, treatment efficacy, presence of adverse effects). Third, the interactions found in our data are only suggestive and our findings require replication in independent populations to confirm their validity. Finally, the characteristics of our participants (individuals of northern European ancestry with moderate-high socioeconomic status) limit our capacity to generalize our conclusions to other populations.

## CONCLUSIONS

In summary, genetic variants in MRP2-*ABCC2* and MDR1-*ABCB1* placental transporters may modulate the association between prenatal exposure to antiseizure drugs and low birth weight in the offspring. To the best of our knowledge, this is the largest gene-drug interaction study on offspring birth weight to date and it was performed in a well-characterized population with genome-wide genotype information. If our findings are confirmed, particularly in studies in which we know exactly which drug the mothers have used during pregnancy, the assessment of these genetic variants may be implemented in clinical settings to support safer use of antiseizure drugs during pregnancy.

## DATA AVAILABILITY STATEMENT

Consent given by the participants does not open for storage of data on an individual level in repositories or journals. Researchers who want access to datasets for replication should apply to. Access to datasets requires approval from the Regional Committee for Medical and Health Research Ethics in Norway and an agreement with MoBa.

## ACKNOWLEDGMENTS

The MoBa Cohort Study is supported by the Norwegian Ministry of Health and Care Services and the Ministry of Education and Research. We thank all the participating families in Norway who take part in this ongoing cohort study, and those who contributed to the recruitment and the infrastructure of the cohort.

We thank the Norwegian Institute of Public Health for generating high-quality genomic data. This research is part of the HARVEST collaboration, supported by the Research Council of Norway (project reference: #229624). We also thank the NORMENT Centre for providing genotype data, funded by the Research Council of Norway (project reference: #223273), South East Norway Health Authority, and Stiftelsen Kristian Gerhard Jebsen. We further thank the Center for Diabetes Research (University of Bergen) for providing genotype data funded by the European Research Council Advanced Grant project SELECTionPREDISPOSED, Stiftelsen Kristian Gerhard Jebsen, the Trond Mohn Foundation, the Research Council of Norway, the Novo Nordisk Foundation, the University of Bergen, and the Western Norway Health Authority.

This work was performed on the TSD (Tjeneste for Sensitive Data) facilities, owned by the University of Oslo, operated, and developed by the TSD service group at the University of Oslo, IT-Department.

This paper does not necessarily reflect the position or policy of the Norwegian Research Council.

## FUNDING

The MoBa Cohort Study is supported by the Norwegian Ministry of Health and Care Services and the Norwegian Ministry of Education and Research. This project received funding from the European Research Council under the European Union’s Horizon 2020 research and innovation program (grant agreement No 947684). This work was also partly supported by the Research Council of Norway through its Centres of Excellence funding scheme, project number 262700 and 223273, and the project “Women’s fertility – an essential component of health and well-being”, number 320656, and co-funded by the European Research Council (grant agreement No 101071773). M.H.H. was supported by the fellowship “*Estancias de movilidad Jose Castillejo dentro del Programa Estatal de Promoción del Talento y su empleabilidad, en el marco del Plan Estatal de lnvestigación Científica y Técnica y de lnnovación 2017-2020”,* from the Spanish government. P.R.N. was supported by the European Research Council (grant agreement No 293574), the Novo Nordic Foundation (#NNF19OC0054741) and the Trond Mohn Foundation (PRECISE-DIA). E.C. and A.Havdahl were supported by the Research Council of Norway (274611) and the South-Eastern Norway Regional Health Authority (project numbers 2020022, 2021045). O.A.A. was supported by the European Research Council (grant agreement No 964874) and the Research Council of Norway (300309, 273291). The funders had no role in the study design; the collection, analysis, and interpretation of data; in the writing of the report; or in the decision to submit the article for publication. Views and opinions expressed in this paper are those of the authors only and do not necessarily reflect those of the funders. Neither the European Union nor the granting authority can be held responsible for them.

## CONFLICTS OF INTEREST

O.A.A. is a consultant for cortechs.ai and has received speaker’s honorarium from Janssen, Lundbeck, and Sunovion unrelated to the current work. The rest of the authors declare that no competing interests exist.

## AUTHORS’ CONTRIBUTION

M.H.H. conceived and designed the study, was responsible for data curation, formal analyses, data interpretation, and drafting the article. J.M.C. critically revised the article and contributed to interpretation of the data. K.H.S. provided support in data analysis, software use, and visualization of results, and critically revised the article. T.K.W.G. was involved in the interpretation of data and critically revised the article. Y.L. contributed to data acquisition and critically revised the article. P.M. contributed to data acquisition and critically revised the article. P.R.N. contributed to data acquisition and critically revised the article. O.A.A. contributed to data acquisition and critically revised the article. E.C. contributed to data acquisition and critically revised the article. A.Havdahl contributed to data acquisition and critically revised the article. E. Molden critically revised the article. S.E.H. contributed to data acquisition, obtained funding, and critically revised the article. K.F. provided support in data analysis and critically revised the article. M.C.M. and A.Hernaez coordinated the project, contributed to data interpretation, and critically revised the article. M.H.H. and A.Hernaez are the guarantors of this study, accept full responsibility for the work and the conduct of the study, had access to the data, and controlled the decision to publish.

## SUPPORTING INFORMATION

### SUPPLEMENTAL MATERIALS

#### Search strategy of genetic variants

All studies that examined the association between the placental transporters of interest and epilepsy or depression were researched. We systematically searched PubMed, Web of Science, pharmacogenetic websites (https://www.pharmgkb.org) and GWAS catalog (https://www.ebi.ac.uk/gwas/home). All databases were searched using the Boolean method with the following terms (1 AND 2 AND 3): 1) “antiseizure” OR “antiepilepsy” OR “anticonvulsants” OR “antidepressant” OR “benzodiazepines”; 2) “polymorphism” OR “genetic polymorphism” OR “genetic variant” OR “pharmacogenetics”; 3) “ATP binding cassette” or “ABCB1” OR “ABCC1” OR “ABCC2” OR “ABCG2” OR “ABCC5” OR “ABCC3”. In addition, the reference and discussions of all pooled articles were carefully scanned for additional publications. We excluded studies: 1) not performed in humans; 2) not published in English; 3) not published in scientific journals; and 4) focused on other drug groups.

### SUPPLEMENTAL TABLES

**Supplemental Table 1.**
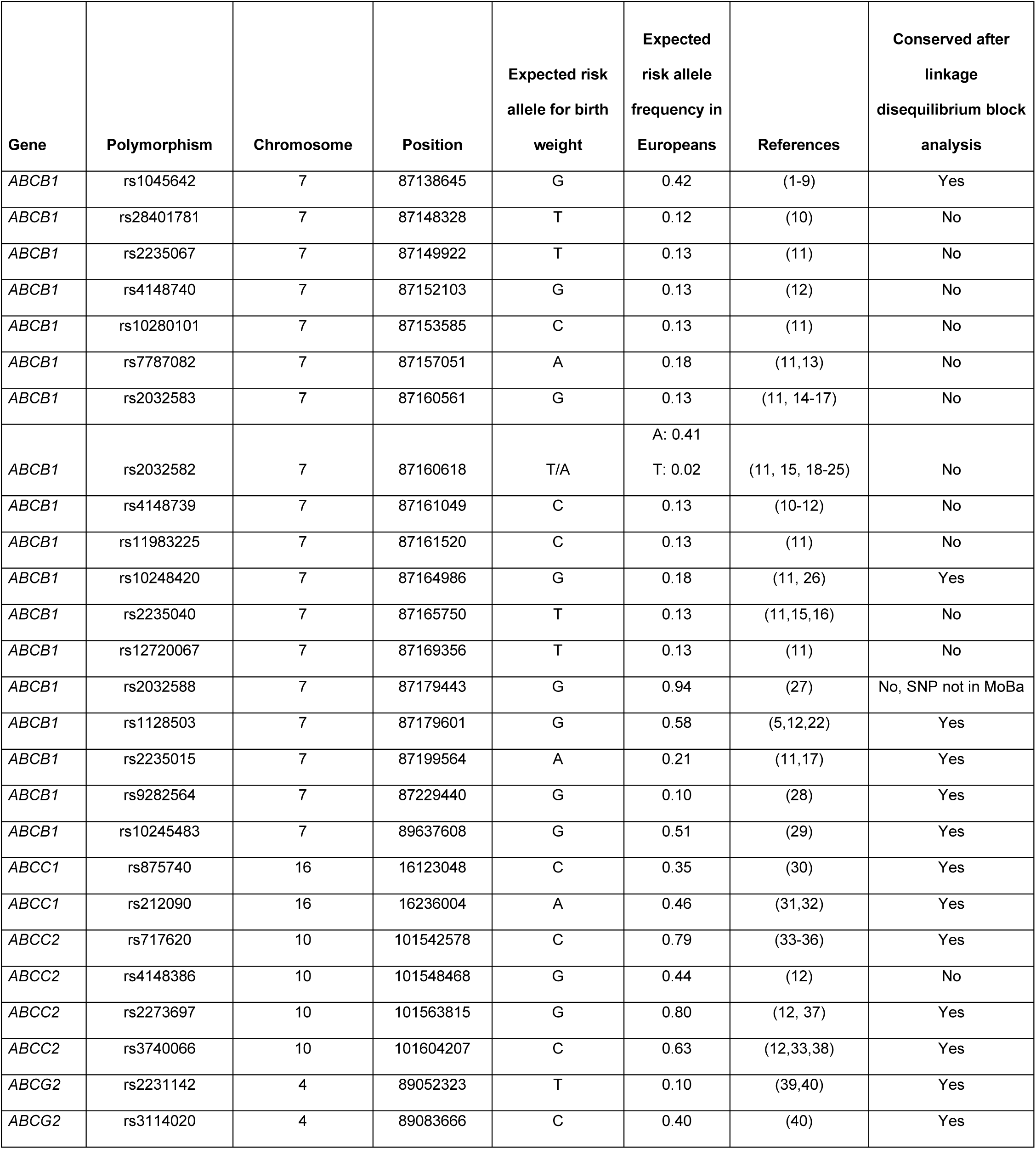
Expected risk allele for birth weight in efflux transporters.

| Gene | Polymorphism | Chromosome | Position | Expected risk allele for birth weight | Expected risk allele frequency in Europeans | References | Conserved after linkage disequilibrium block analysis |
| --- | --- | --- | --- | --- | --- | --- | --- |
| *ABCB1* | rs1045642 | 7 | 87138645 | G | 0.42 | (1-9) | Yes |
| *ABCB1* | rs28401781 | 7 | 87148328 | T | 0.12 | (10) | No |
| *ABCB1* | rs2235067 | 7 | 87149922 | T | 0.13 | (11) | No |
| *ABCB1* | rs4148740 | 7 | 87152103 | G | 0.13 | (12) | No |
| *ABCB1* | rs10280101 | 7 | 87153585 | C | 0.13 | (11) | No |
| *ABCB1* | rs7787082 | 7 | 87157051 | A | 0.18 | (11,13) | No |
| *ABCB1* | rs2032583 | 7 | 87160561 | G | 0.13 | (11, 14-17) | No |
| *ABCB1* | rs2032582 | 7 | 87160618 | T/A | A: 0.41<br>T: 0.02 | (11, 15, 18-25) | No |
| *ABCB1* | rs4148739 | 7 | 87161049 | C | 0.13 | (10-12) | No |
| *ABCB1* | rs11983225 | 7 | 87161520 | C | 0.13 | (11) | No |
| *ABCB1* | rs10248420 | 7 | 87164986 | G | 0.18 | (11, 26) | Yes |
| *ABCB1* | rs2235040 | 7 | 87165750 | T | 0.13 | (11,15,16) | No |
| *ABCB1* | rs12720067 | 7 | 87169356 | T | 0.13 | (11) | No |
| *ABCB1* | rs2032588 | 7 | 87179443 | G | 0.94 | (27) | No, SNP not in MoBa |
| *ABCB1* | rs1128503 | 7 | 87179601 | G | 0.58 | (5,12,22) | Yes |
| *ABCB1* | rs2235015 | 7 | 87199564 | A | 0.21 | (11,17) | Yes |
| *ABCB1* | rs9282564 | 7 | 87229440 | G | 0.10 | (28) | Yes |
| *ABCB1* | rs10245483 | 7 | 89637608 | G | 0.51 | (29) | Yes |
| *ABCC1* | rs875740 | 16 | 16123048 | C | 0.35 | (30) | Yes |
| *ABCC1* | rs212090 | 16 | 16236004 | A | 0.46 | (31,32) | Yes |
| *ABCC2* | rs717620 | 10 | 101542578 | C | 0.79 | (33-36) | Yes |
| *ABCC2* | rs4148386 | 10 | 101548468 | G | 0.44 | (12) | No |
| *ABCC2* | rs2273697 | 10 | 101563815 | G | 0.80 | (12, 37) | Yes |
| *ABCC2* | rs3740066 | 10 | 101604207 | C | 0.63 | (12,33,38) | Yes |
| *ABCG2* | rs2231142 | 4 | 89052323 | T | 0.10 | (39,40) | Yes |
| *ABCG2* | rs3114020 | 4 | 89083666 | C | 0.40 | (40) | Yes |

**Supplemental Table 2.** The articles detail the development of the expected risk allele for the transporter score.

| Gene | Polymorphism | Drug | Study participants | Effect |  | P-value | Expected risk allele in the publication | Risk allele for birth weight | Reference |
| --- | --- | --- | --- | --- | --- | --- | --- | --- | --- |
| *ABCB1* | rs1045642 | All antiseizure drugs | 114 drug-resistant epilepsy patients, 213 seizure-free patients and 287 controls (Taiwan) | Efficacy | The C allele and the CC genotype were associated with drug resistance relative to the T allele and the TT genotype. | 0.001 | T | G | 1 |
| *ABCB1* |  | All antiseizure drugs | 746 patients with epilepsy and 179 controls (China) | Efficacy | The TT genotype was associated with drug resistance | 0.0009 | G |  | 2 |
| *ABCB1* |  | All antiseizure drugs | 220 patients with epilepsy and 220 controls (India) | Efficacy | The TT genotype was associated with drug resistance relative to the CC genotype. | 0.001 | G |  | 3 |
| *ABCB1* |  | Carbamazepine | 3,293 patients with epilepsy. Meta analysis | Pharmacokinetics/<br>Efficacy | The T allele was associated with lower concentrations, lower absorption, and resistance to the drug activity. | 0.004 | T |  | 4 |
| *ABCB1* |  | Carbamazepine | 2,126 patients with epilepsy. Meta-analysis | Efficacy | The T allele was associated to decreased plasma concentrations of the drug. | <0.05 | G |  | 5 |
| *ABCB1* |  | Agomelatine | 28 healthy volunteers (Spain) | Pharmacokinetics | The TT genotype was associated with lower concentrations or lower absorption of the drug. | 0.047 | G |  | 6 |
| *ABCB1* |  | All antiseizure drugs | 8,716 patients with epilepsy, 4,037 drug-resistant epilepsy patients and 4,679 drug-responsive patients. Meta analysis | Efficacy | The T allele was associated with higher risk of drug resistance in the overall population and in Caucasians. | 0.006 | G |  | 7 |
| *ABCB1* |  | Carbamazepine | In vitro | Impact of T allele on the sensitivity, intracellular accumulation, and transepithelial permeability of antiseizure drugs | The recombinant T allele_cells showed higher resistance to carbamazepine compared with C allele_cells in the cytotoxicity assay. The intracellular accumulation of carbamazepine was significantly decreased in cells transfecting with recombinant T allele when compared with recombinant C allele_cells. | 0.01 | G |  | 8 |
| *ABCB1* |  | Carbamazepine | 210 patients with epilepsy (China) | Pharmacokinetics | The GG genotype was associated with increased concentrations of carbamazepine relative to the AG genotype. | 0.001 | G |  | 9 |
| *ABCB1* | rs10248420 | All antidepressants | 256 patients with epilepsy (Europe) | Efficacy | The G allele was associated with increased likelihood of remission when treated with amitriptyline, citalopram, paroxetine or venlafaxine relative to the A allele. | 0.0095 | G | G | 11 |
| *ABCB1* |  | Olanzapine | 80 healthy volunteers (Spain) | Efficacy | The A allele was associated with decreased drug response compared to the G allele | 0.047 | G |  | 26 |
| *ABCB1* | rs1128503 | All antiseizure drugs | 90 patients with epilepsy (USA) | Pharmacokinetics | AA and AG genotypes were associated with increased clearance of carbamazepine relative to the GG genotype. | 0.036 | G | G | 12 |
| *ABCB1* |  | Lamotrigine | 222 patients with epilepsy (Croatia) | Pharmacokinetics | The genotype CC was associated with higher lamotrigine concentrations compared to CT and TT genotypes. | 0.021 | G |  | 22 |
| *ABCB1* |  | Carbamazepine | 2,126 patients with epilepsy. Meta-analysis | Efficacy | The CC genotype was associated with lower resistance to carbamazepine. | 0.03 | G |  | 5 |
| *ABCB1* | rs2235015 | All antidepressants | 339 patients with depression (European) | Efficacy | The A allele was associated with higher remission for P-glycoprotein antidepressants, citalopram, paroxetine, amitriptyline, and venlafaxine, relative to the C allele. | 0.025 | A | A | 11 |
| ABCB1 |  | All antidepressants | 73 patients with depressive disorder (Germany) | Efficacy | The A allele was associated with increased likelihood of remission when treated with amitriptyline, citalopram, paroxetine or venlafaxine, relative to the C allele. | 0.030 | A |  | 17 |
| ABCB1 | rs9282564 | Paroxetine | 71 psychiatric patients (Switzerland) | Efficacy | The G allele was associated with greater clinical response. | 0.043 | G | G | 28 |
| ABCB1 | rs10245483 | All antidepressants | 683 patients with major depressive disorder (multisite) | Efficacy | The T allele was associated with less remission. | 0.001 | G | G | 29 |
| ABCC1 | rs875740 | All antiseizure drugs | 199 patients from the Mayo Clinic Bipolar Disorder Biobank GWAS (USA) | Efficacy | The C allele was associated with better treatment response. | 0.0089 | C | C | 30 |
| ABCC1 | rs212090 | Clozapine | 137 medicated patients (Australia) | Adverse effects | The A allele was associated with higher diastolic blood pressure (adverse effect) | 0.01 | A | A | 31 |
| ABCC1 |  | All antidepressants | 148 patients with major depressive disorder (Australia) | Efficacy | The A allele was included int the pharmacogenetic guide group and associated with greater remission | 0.0001 | A |  | 32 |
| ABCC2 | rs717620 | All antiseizure drugs | 537 patients with epilepsy (217 drug resistant patients and 320 drug responders) (China) | Efficacy | The TT genotype was associated with drug resistance | 0.001 | C | C | 33 |
| ABCC2 |  | All antiseizure drugs | 4,300 patients with epilepsy (2,261 drug-resistant patients and 2,039 drug-responsive patients). Meta-analysis | Efficacy | The T allele was associated with drug resistance | 0.0006 | C |  | 34 |
| ABCC2 |  | All antiseizure drugs | 254 patients with epilepsy (104 drug-resistant and 150 drug-responsive) (China) | Efficacy | The TT genotype was associated with drug resistance, relative to the CC genotype | 0.001 | C |  | 35 |
| ABCC2 |  | All antiseizure drugs | 1,842 patients with epilepsy. Meta-analysis | Efficacy | The T allele was associated with drug resistance. | 0.002 | C |  | 36 |
| ABCC2 | rs2273697 | All antiseizure drugs | 90 patients with epilepsy (USA) | Pharmacokinetics | AA and AG genotypes were associated with higher drug clearance relative to the GG genotype. | <0.05 | G | G | 12 |
| ABCC2 |  | All antiseizure drugs | 453 patients with epilepsy (China) | Efficacy | The A allele A was associated with drug resistance. | 0.001 | G |  | 37 |
| ABCC2 | rs3740066 | All antiseizure drugs | 90 patients with epilepsy (USA) | Pharmacokinetics | The G allele was associated with lower carbamazepine-10, 11-epoxide (metabolite): carbamazepine ratio. | 0.008 | C | C | 12 |
| ABCC2 |  | All antiseizure drugs | 537 patients with epilepsy (217 drug resistant patients and 320 drug responders) (China) | Efficacy | CT and TT genotypes were associated with drug resistance relative to CC genotype. | 0.038 | C |  | 33 |
| ABCC2 |  | No drug | In vitro study | In vitro function of the transporter | The T allele was associated with a decreased function of the transporter. | 0.017 | C |  | 38 |
| ABCG2 | rs2231142 | Lamotrigine | 112 patients with epilepsy (China) | Pharmacokinetics | The TT genotype was associated with increased concentrations of lamotrigine relative to GG and GT genotypes. | 0.015 | T | T | 39 |
| ABCG2 |  | Lamotrigine | 140 patients with epilepsy (China) | Pharmacokinetics | CA and AA genotypes were associated with higher concentrations of lamotrigine relative to the CC genotype. | <0.05 | T |  | 40 |
| ABCG2 | rs3114020 | Lamotrigine | 140 patients with epilepsy (China) | Pharmacokinetics | CC and CT genotypes were associated with increased concentrations of lamotrigine relative to the TT genotype. | 0.01 | C | C | 40 |

**Supplemental Table 3.** STROBE checklist for cohort studies.

|  | Item No | Recommendation | Page No |
| --- | --- | --- | --- |
| Title and abstract |  |  |  |
| Title and abstract | 1 | (a) Indicate the study's design with a commonly used term in the title or the abstract | 1 |
|  |  | (b) Provide in the abstract an informative and balanced summary of what was done and what was found | 3-4 |
| Introduction |  |  |  |
| Background/<br>rationale | 2 | Explain the scientific background and rationale for the investigation being reported | 6-7 |
| Objectives | 3 | State specific objectives, including any pre-specified hypotheses | 6-7 |
| Methods |  |  |  |
| Study design | 4 | Present key elements of study design early in the paper | 6 |
| Setting | 5 | Describe the setting, locations, and relevant dates, including periods of recruitment, exposure, follow-up, and data collection | 8 |
| Participants | 6 | (a) Give the eligibility criteria, and the sources and methods of selection of participants. Describe methods of follow-up | 8-9 |
|  |  | (b) For matched studies, give matching criteria and number of exposed and unexposed | - |
| Variables | 7 | Clearly define all outcomes, exposures, predictors, potential confounders, and effect modifiers. Give diagnostic criteria, if applicable | 9-11 |
| Data sources/<br>measurement | 8 | For each variable of interest, give sources of data and details of methods of assessment (measurement). Describe comparability of assessment methods if there is more than one group | 8-11 |
| Bias | 9 | Describe any efforts to address potential sources of bias | 9-11 |
| Study size | 10 | Explain how the study size was arrived at | Fig.1 |
| Quantitative variables | 11 | Explain how quantitative variables were handled in the analyses. If applicable, describe which groupings were chosen and why | 10 |
| Statistical methods | 12 | (a) Describe all statistical methods, including those used to control for confounding | 10-11 |
|  |  | (b) Describe any methods used to examine subgroups and interactions | 10-11 |
|  |  | (c) Explain how missing data were addressed | - |
|  |  | (d) If applicable, explain how loss to follow-up was addressed | - |
|  |  | (e) Describe any sensitivity analyses | 11 |
| Results |  |  |  |
| Participants | 13* | (a) Report numbers of individuals at each stage of study—e.g. numbers potentially eligible, examined for eligibility, confirmed eligible, included in the study, completing follow-up, and analyzed | Fig. 1 |
|  |  | (b) Give reasons for non-participation at each stage |  |
|  |  | (c) Consider use of a flow diagram |  |
| Descriptive data | 14* | (a) Give characteristics of study participants (e.g. demographic, clinical, social) and information on exposures and potential confounders | 11, Table 1 |
|  |  | (b) Indicate number of participants with missing data for each variable of interest | - |
|  |  | (c) Summarize follow-up time (e.g., average and total amount) | - |
| Outcome data | 15 | Report numbers of outcome events or summary measures over time | 14-18 |
| Main results | 16 | (a) Give unadjusted estimates and, if applicable, confounder-adjusted estimates and their precision (e.g., 95% confidence interval). Make clear which confounders were adjusted for and why they were included | 14-18<br>Figures 2,3 |
|  |  | (b) Report category boundaries when continuous variables were categorized | 14-18<br>Figures 2,3 |
|  |  | (c) If relevant, consider translating estimates of relative risk into absolute risk for a meaningful time period | - |
| Other analyses | 17 | Report other analyses done—e.g. analyses of subgroups and interactions, and sensitivity analyses | 18,<br>Supplemental figures 2-5 |
| Discussion |  |  |  |
| Key results | 18 | Summarize key results with reference to study objectives | 19 |
| Limitations | 19 | Discuss limitations of the study, taking into account sources of potential bias or imprecision. Discuss both direction and magnitude of any potential bias | 20 |
| Interpretation | 20 | Give a cautious overall interpretation of results considering objectives, limitations, multiplicity of analyses, results from similar studies, and other relevant evidence | 19-20 |
| Generalizability | 21 | Discuss the generalizability (external validity) of the study results | 20 |
| Other information |  |  |  |
| Funding | 22 | Give the source of funding and the role of the funders for the present study and, if applicable, for the original study on which the present article is based | 23 |

### SUPPLEMENTAL RESULTS

**Supplemental Figure 1.**
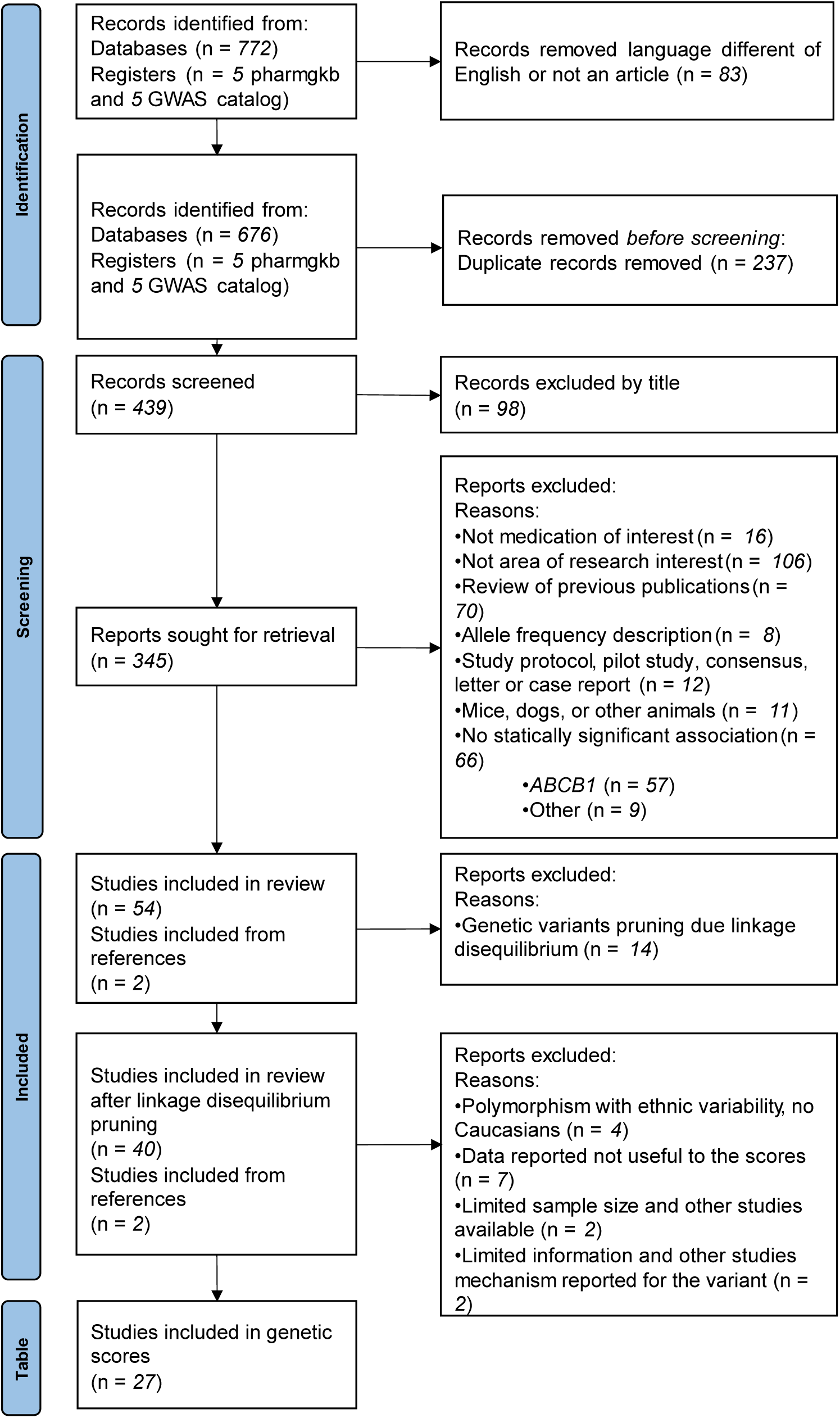
Flow chart of the systematic literature review of genetic variants on placental efflux transporters.

**Supplemental Figure 2.**
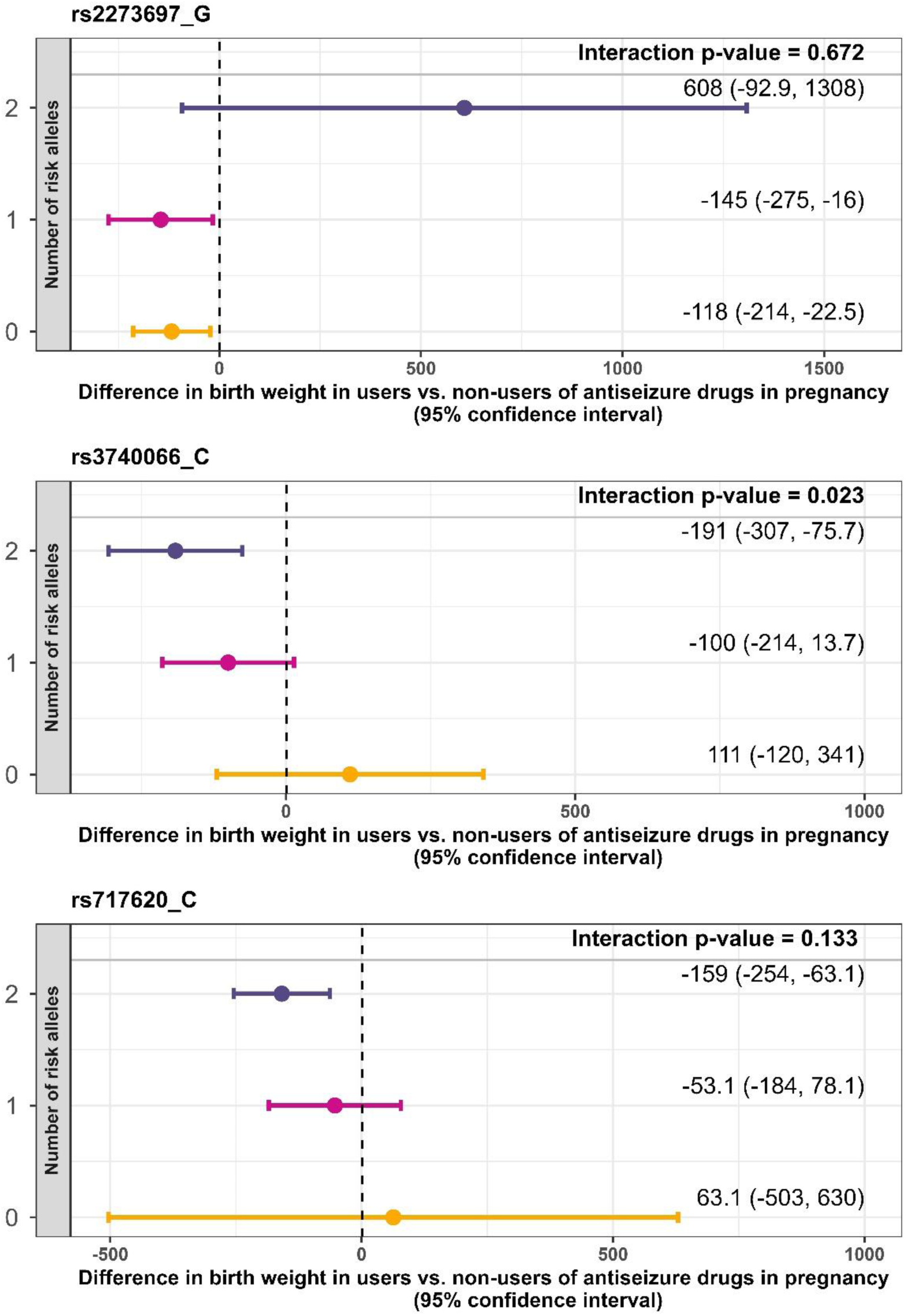
Differences in birth weight associated with pregnancy use of antiseizure drugs due to *ABCC2* individual genetic variants in the offspring.

**Supplemental Figure 3.**
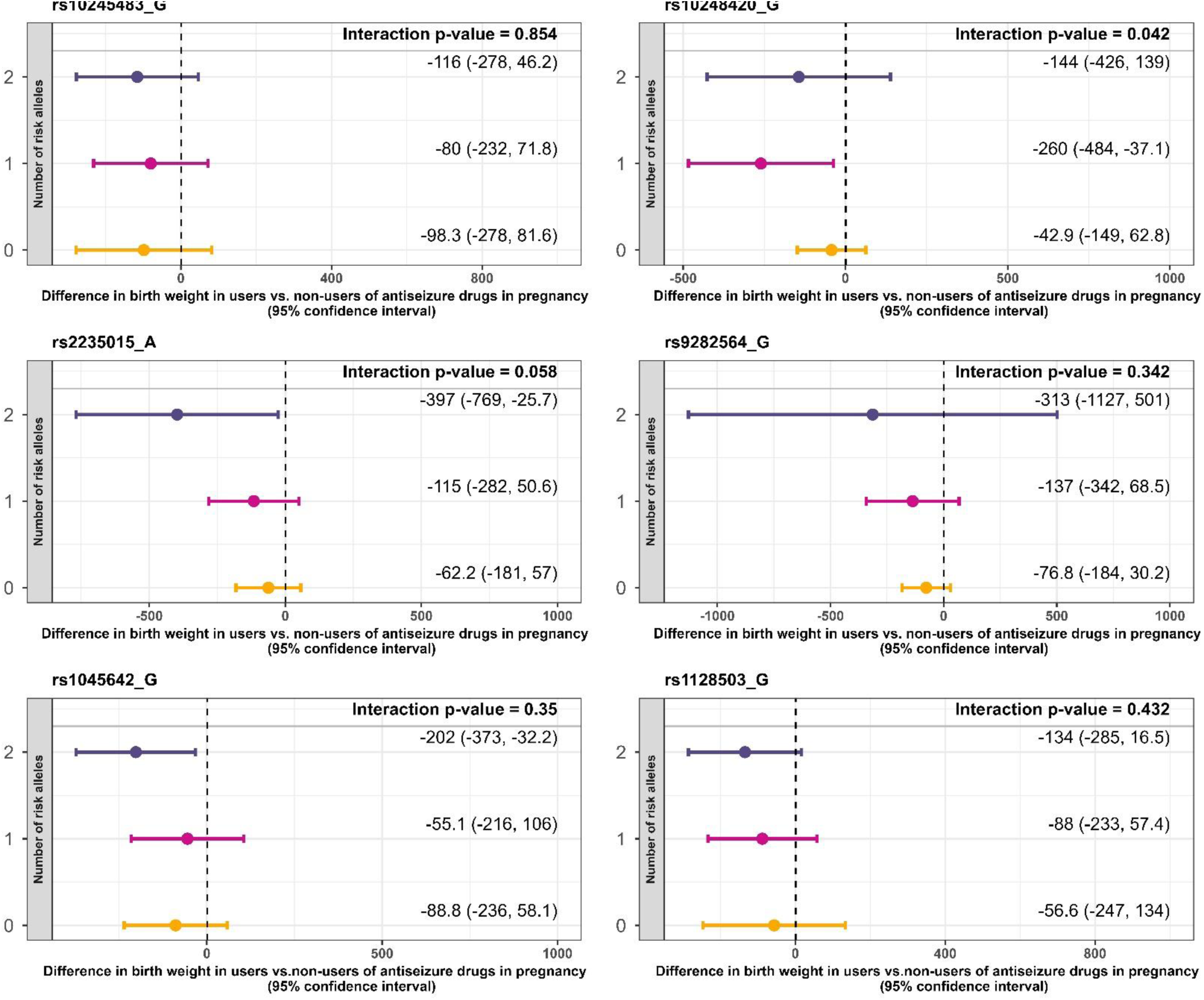
Differences in birth weight associated with pregnancy use of antiseizure drugs due to *ABCB1* individual genetic variants in the mother.

**Supplemental Figure 4.**
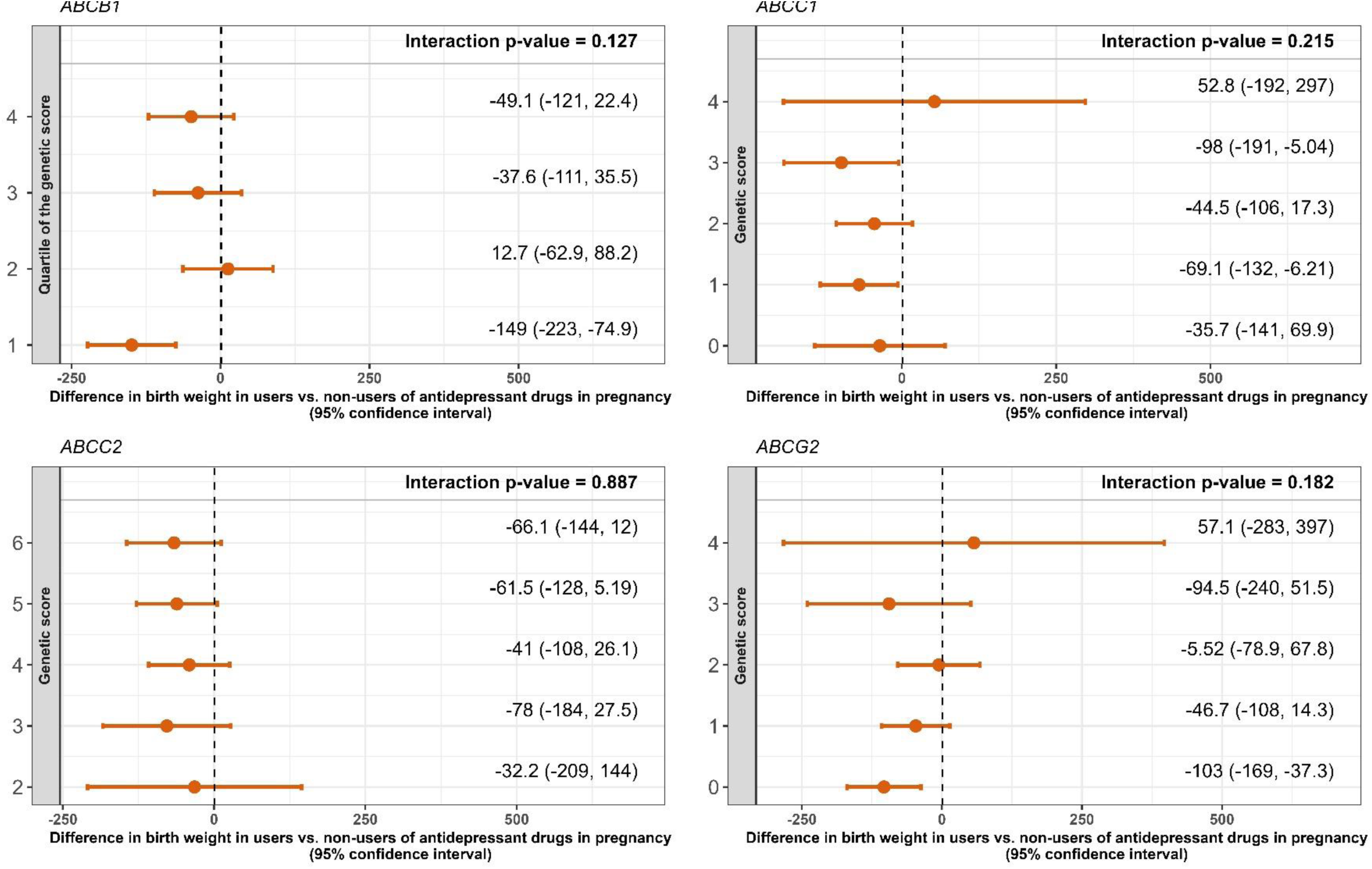
Differences in birth weight associated with pregnancy use of antidepressant medication and placental transporter genetic scores in the offspring.

**Supplemental Figure 5.**
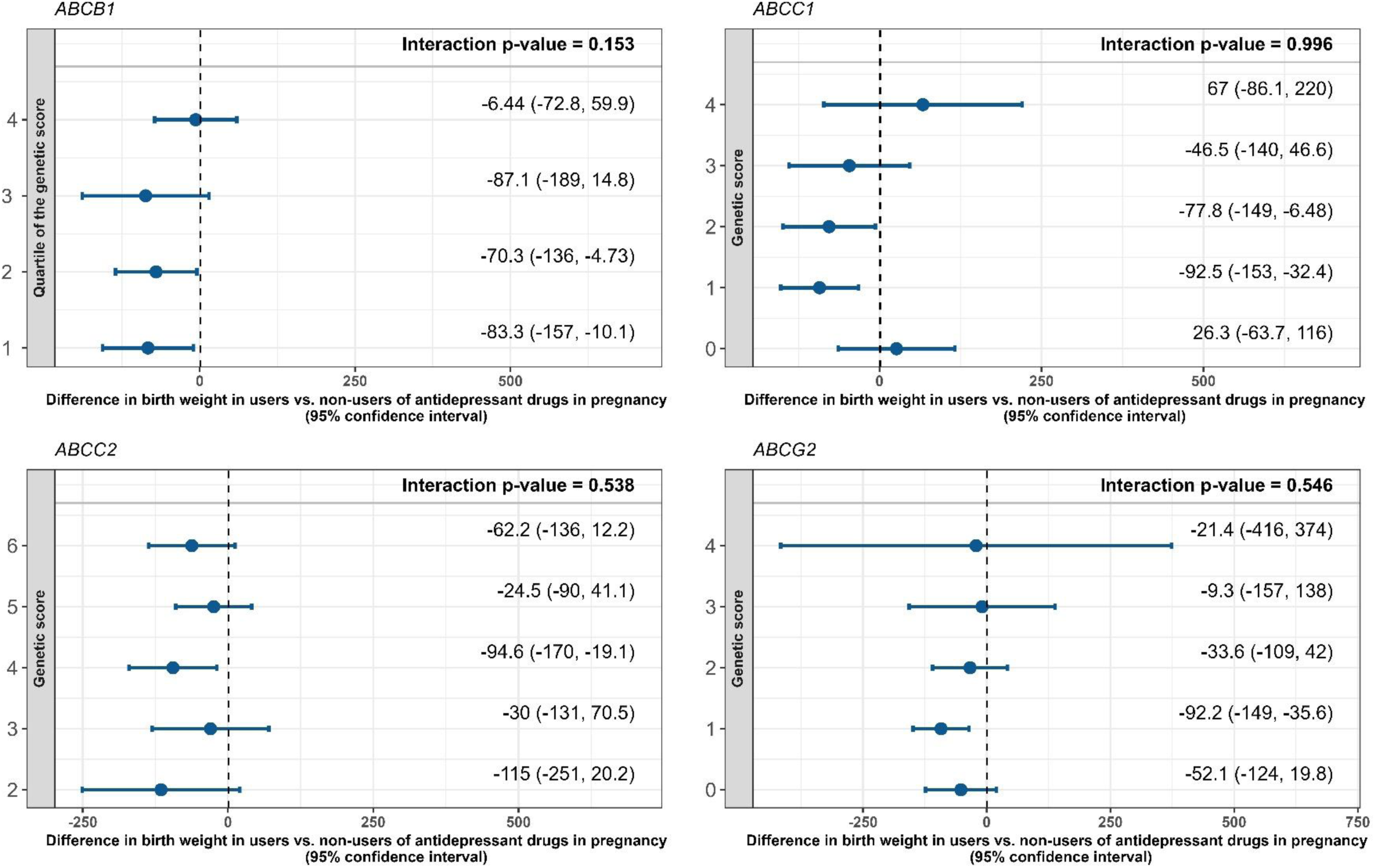
Differences in birth weight associated with pregnancy use of antidepressant medication and placental transporter genetic scores in the mother.

